# Quantitative observational evidence of indirect herd benefits from COVID-19 vaccination or prior infection on SARS-CoV-2 infections and COVID-19 deaths: a population-based retrospective cohort study in Ontario, Canada

**DOI:** 10.1101/2025.01.30.25321209

**Authors:** Linwei Wang, Sarah Swayze, Arjumand Siddiqi, Stefan D. Baral, Beate Sander, Hind Sbihi, Jeffrey C. Kwong, Sharmistha Mishra

## Abstract

**Background:** Empirical evidence on the indirect herd benefits of COVID-19 vaccination and/or prior infection is limited. We aimed to examine how area-level immunity interacts with individual-level immunity to affect COVID-19 diagnoses and deaths.

**Methods:** Ontario residents aged ≥18 years were followed from August-01-2021 to January-30-2022. Individual-level immunity was defined as received a primary series of COVID-19 vaccines or a positive SARS-CoV-2 test in the past 165 days. Area-level immunity was determined based on the proportion of immune individuals in an individual’s residing area. We used logistic regression and cause-specific hazard models to examine the relationship between immunity and COVID-19 diagnosis, and between immunity and COVID-19 death, respectively. We included an interaction term between individual-level and area-level immunity in each model.

**Results:** Of 11,122,816 adults, 7,518,015 (67.6%) were immune at baseline. After accounting for individual-level demographics, baseline health, and area-level social determinants of health, area-level immunity (highest vs. lowest quintiles) was associated with lower odds of COVID-19 diagnosis; the association was larger among non-immune (odds ratios [95% confidence interval]: 0.72 [0.70, 0.75]) than immune individuals (0.93 [0.90, 0.96]). Higher area-level immunity (highest vs. lowest quintiles) was also associated with lower hazard of COVID-19 death among non-immune individuals (hazard ratio: 0.77 [0.60, 1.00]).

**Conclusions:** Our study provides observational evidence supporting the herd benefits of vaccination or prior infection on SARS-CoV-2 infections and COVID-19 deaths. Findings reinforce the need for high vaccination coverage to protect vaccinated and unvaccinated populations, while providing insights for interpreting vaccine effectiveness estimates in the context of herd immunity.

**Main points:** Using area-level immunity coverage as a proxy for herd immunity, our study demonstrates observational evidence of indirect herd benefits from vaccination and/or prior infection on SARS-CoV-2 infections and COVID-19 deaths. Moreover, herd benefits are greater among non-immune individuals.

## Introduction

Vaccines can provide direct protection against infection and against morbidity and mortality(1,2). These mechanisms are key to the design, implementation, and evaluation of COVID-19 vaccination programs. COVID-19 vaccines have been shown in clinical trials and observational studies to provide direct benefits to those immunized by lowering the risk of acquiring infection and reducing disease severity if infected(3). Indirect benefits can arise from a decrease in infectiousness among vaccinated persons if they become infected, leading to fewer infections among their contacts(4), as evidenced through observational studies that measured secondary attack rates in household contacts of vaccinated persons(5,6). Indirect benefits can also arise from herd immunity (hereafter referred to as “indirect herd benefits”) because there are fewer susceptible persons through vaccination and/or prior infection, thereby reducing the overall spread of disease(7,8).

The indirect herd benefits of vaccination and/or prior infection are traditionally quantified using mathematical models that simulate the transmission dynamics of a pathogen(9–11). Transmission dynamics models are explicitly set up to capture feedback loops between prevalent and incident infections, and thus, can be designed to estimate indirect herd benefits(9–11). For example, Scutt et al. used a Susceptible-Infected-Recovered model of SARS-CoV-2 transmission dynamics and demonstrated that under certain epidemic conditions, a larger number of deaths could be prevented from indirect herd benefits of vaccination than from direct benefits(10). However, there remains a gap in empirical evidence on the indirect herd benefits of COVID-19 vaccination and/or prior infection.

There are some empirical data on the indirect herd benefits of vaccination for other infectious diseases(12,13). Chief among them is a cluster randomized trial which showed that high vaccination coverage among children and adolescents in Hutterite communities reduced influenza infections among unvaccinated residents of those communities(12). However, randomized trials to measure the indirect herd benefits of COVID-19 vaccination would be unethical. Alternatives may include leveraging observational data to investigate how area-level vaccination coverage influences individual risk of infection and/or disease progression. One study from King County, US, compared the risk of pertussis among non-fully vaccinated children residing in census tracts with the highest quartile pertussis vaccination coverage with that in the lowest quartile vaccination coverage to measure the indirect vaccine benefits(14). Area-level measures, which reflect individuals’ social network and contact patterns, have been used in other studies to assess the impact of structural and environmental factors on health outcomes(15,16). Individuals’ own immunity (acquired from either vaccination or prior infection) and the level of immunity of their contact network (captured by the area-level immunity coverage of where an individual lives) could interplay to shape individual risk of infection and/or disease progression.

Using population-based observational data from adult residents in Ontario, Canada, we aimed to examine how area-level immunity coverage interacts with individual-level immunity and their associations with COVID-19 diagnoses and deaths.

## Methods

### Study population and measures

We used data from a population-based retrospective cohort, which included community-dwelling individuals aged 18 years and older residing in Ontario as of August 1, 2021, who were followed to January 30, 2022. Individuals were identified using Ontario’s Registered Persons Database, including those with provincial health insurance and excluding long-term care home residents. Details of the cohort have been described in previous studies(17,18). Our study period captured the fourth (Delta dominant) and the majority of the fifth (Omicron B.1.1.529 dominant) waves of the regional COVID-19 pandemic, a time when vaccination became prevalent (to allow for a proportion of our study population to gain immunity from either vaccination and/or prior infection). We followed the ‘RECORD’ reporting guideline (**Appendix Text 1**).

Our two outcomes of interest were COVID-19 diagnosis (defined as having a positive SARS-CoV-2 test), and COVID-19 death (defined as death within 30 days following or 7 days prior to a positive SARS-CoV-2 test)(17). We ascertained outcomes using records from Ontario’s COVID-19 surveillance database (Public Health Case and Contact Management Solution), Ontario Laboratories Information System, and Ontario’s Registered Persons Database.

Our exposures of interest were individual-level immunity and area-level immunity. Individual-level immunity was determined based on an individual’s vaccination status and prior COVID-19 diagnosis as of August 1, 2021, classified as either being immune (received ≥ 1 dose of the Johnson-Johnson vaccine or ≥ 2 doses of other approved vaccines or had a positive SARS-CoV-2 test, in the past 165 days(19,20)) or non-immune. Vaccination status was obtained from COVaxON, Ontario’s COVID-19 vaccination registry data. We determined vaccination status and diagnosis in the past 165 days to account for potential immune waning(19). Area-level immunity was determined based on the immunity coverage of an individual’s residing forward sortation area (FSA) representing an area with a median of 21,722 (interquartile range: 13,082 to 34,288) residents(21). We defined area-level immunity at the FSA-level to capture a proxy of an individual’s contact network. Previous research on contact patterns in Ontario has shown that individual mobility within FSAs is more frequent than mobility between FSAs(22). Immunity coverage for each FSA was calculated as the proportion of the population who is immune (via vaccination or prior diagnosis as defined above) within that FSA. FSAs were then ranked by immunity coverage and categorized into quintiles (quintile 1 comprised areas with the lowest immunity coverage), with weights adjusted for population size.

We considered the covariates listed in **Figure 1** as potential confounders based on prior literature, including individual-level demographics and baseline health, geographical residence, and area-level social determinants of health(17,18). Details of covariates and their data sources are provided in **Figure 1**, and **Table 1**, and described in prior studies(17,18).

**Figure 1.**
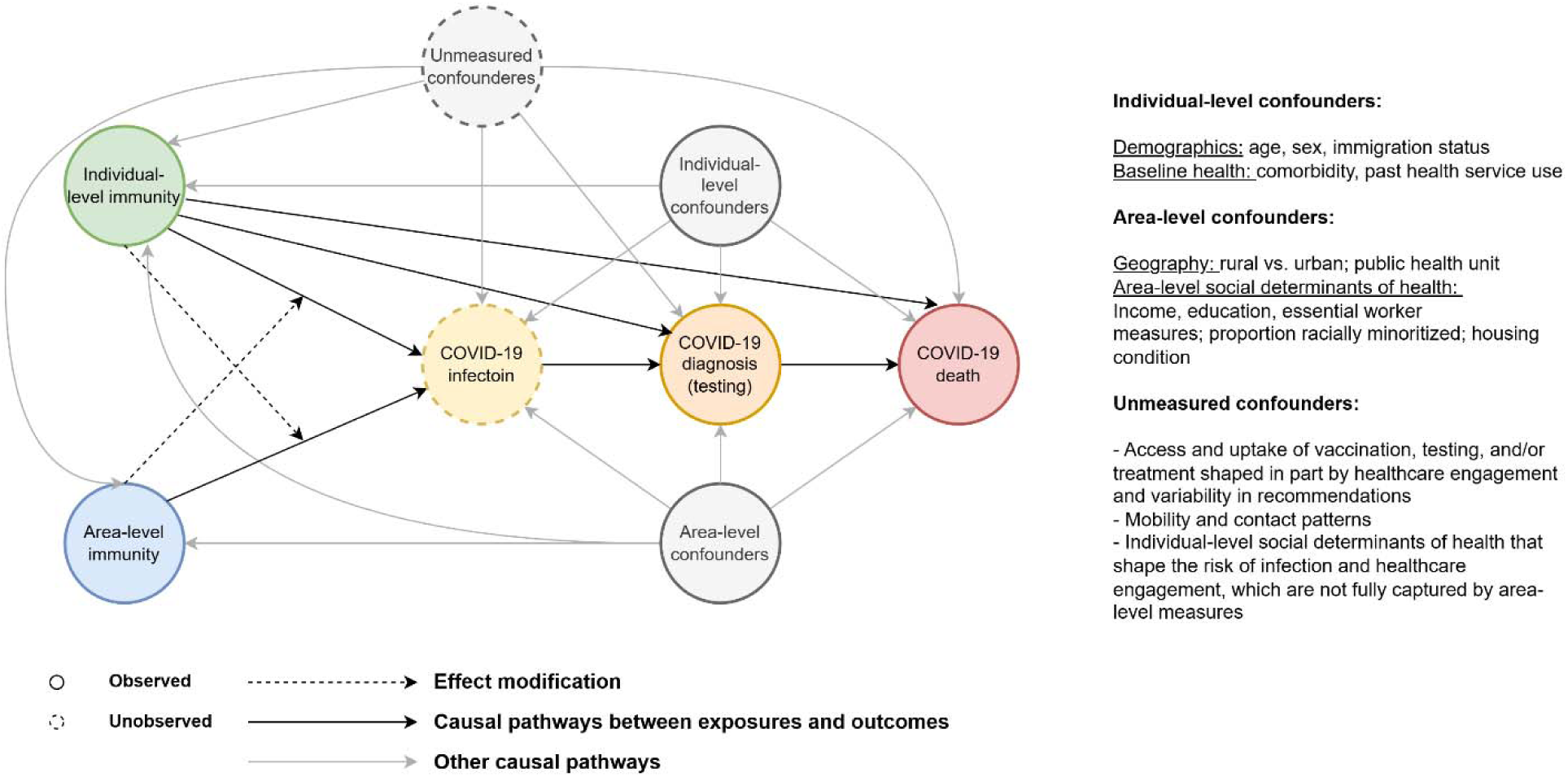
Directed acyclic graphs representing the effects of individual-level immunity and area-level immunity on COVID-19 diagnosis and death, and their effect modification on each other. Individual-level confounders include demographics (age, sex, immigration status), baseline health (comorbidity, past health service use). Area-level confounders include geography (rural vs. urban, public health unit), and area-level social determinants of health (income, education, essential worker measures, proportion racially minoritized, housing condition). Unmeasured confounders may include access and uptake of vaccination, testing, and/or treatment shaped in part by healthcare engagement and variability in recommendations; mobility and contact patterns which affect herd immunity threshold; individual-level social determinants of health that shape the risk of infection and healthcare engagement, which are not fully captured by area-level measures.

**Table 1.**
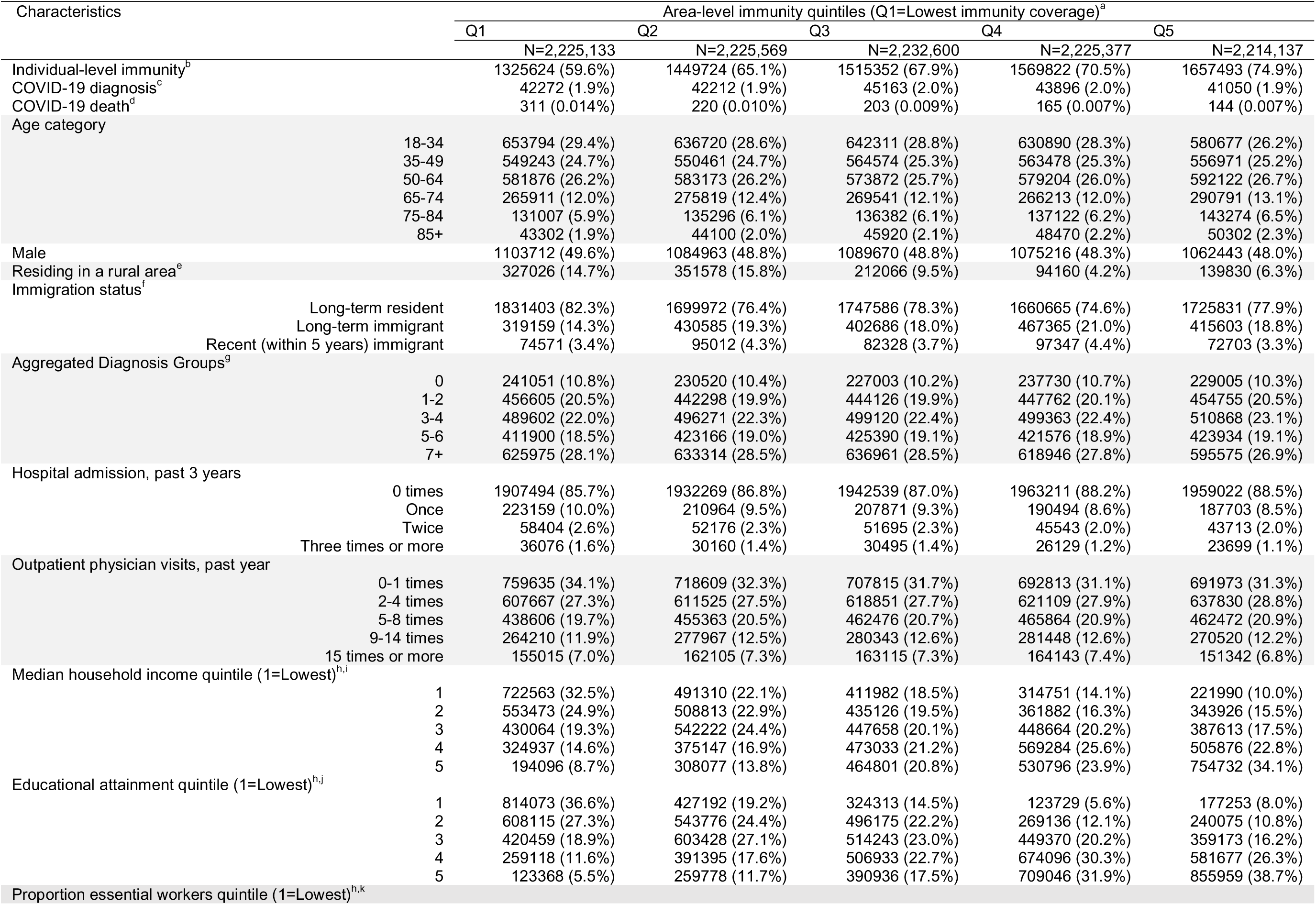
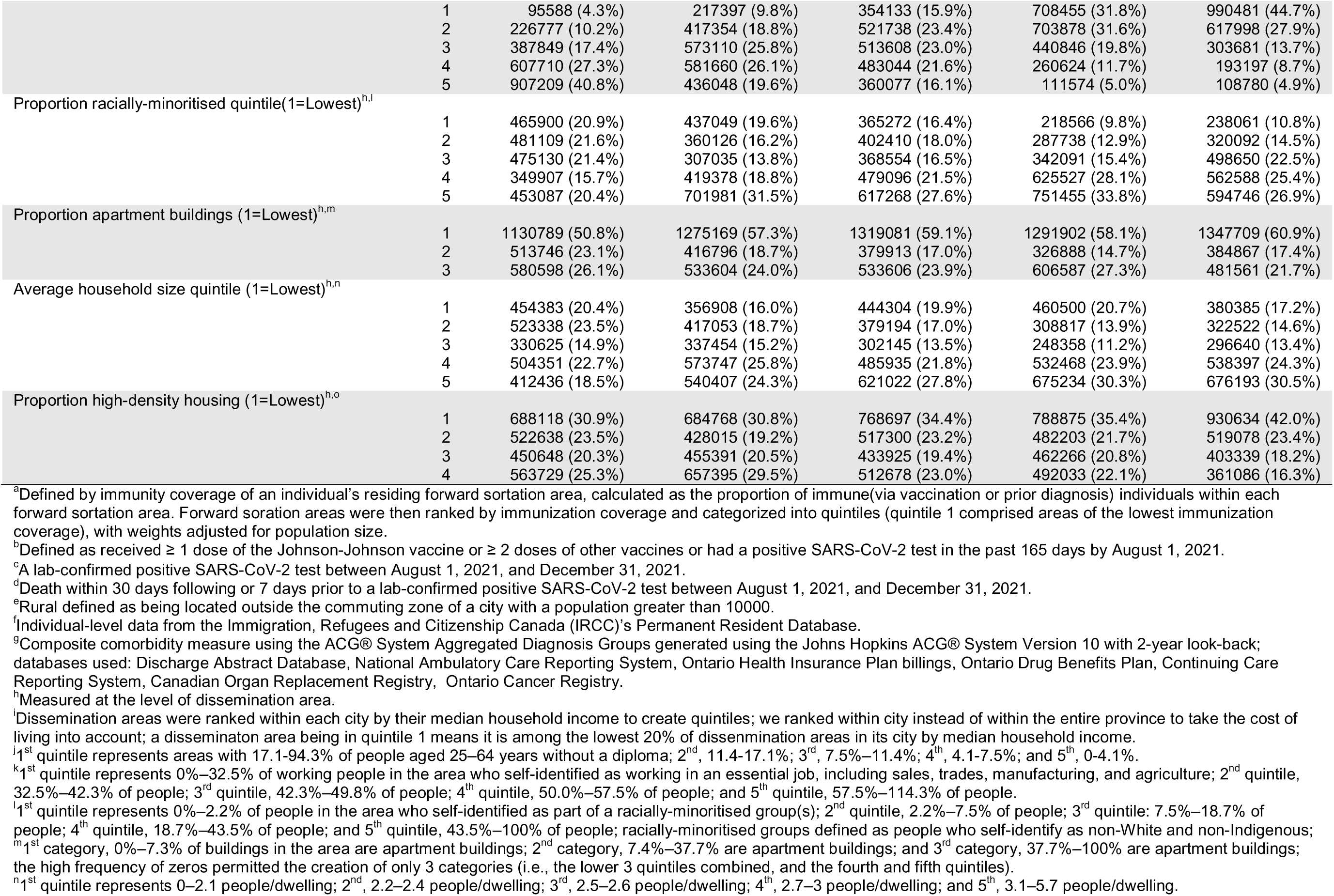
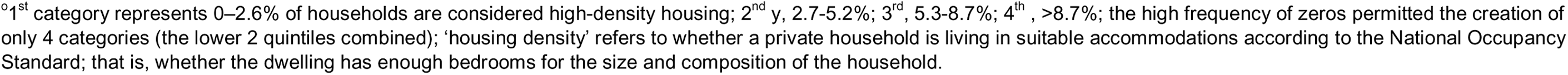
Characteristics of community-dwelling adults in Ontario stratified by area-level immunity quintiles (N=11,122,816).

All data sets were linked using unique encoded identifiers and analyzed at ICES(23). The use of the data in this project is authorized under section 45 of Ontario’s Personal Health Information Protection Act and does not require review by a Research Ethics Board.

### Statistical analyses

We hypothesized that if vaccination and prior infection provided indirect benefits through herd immunity, area-level immunity would be independently associated with COVID-19 diagnosis and death, even after adjusting for individual-level immunity and confounders (depicted using a directed acyclic graph (**Figure 1**)). Moreover, we hypothesized that there would be an interaction between individual-level and area-level immunity, such that the effect of area-level immunity could vary by individual-level immunity, and vice versa. The interaction could be due to immune saturation - where a critical threshold of immunity is reached in the population, limiting the impact of additional immunity on disease transmission(8) (**Figure 1**).

To test our hypotheses, we used logistic regression models to examine the relationship between immunity (both individual-level and area-level) and COVID-19 diagnosis, and cause-specific hazard models(24) to examine the relationship between immunity and COVID-19 death. We included an interaction term between individual-level and area-level immunity in each model and tested for its statistical significance. We assessed the unadjusted relationships to show the observed patterns and those adjusted for confounders to test our hypotheses.

We conducted several sensitivity analyses. To explore whether geographic scale influences the relationship between area-level immunity and COVID-19 outcomes, we repeated our analyses using the area-level immunity measured at the level of dissemination area (representing 400-700 residents, thus smaller than FSA). Immune protection acquired from prior infection might differ from vaccination(25). As such, we repeated our analyses considering immunity from vaccination only, regardless of prior diagnosis.

## Results

Of 11,122,816 adults included in our analyses, 7,518,015 (67.6%) individuals were either vaccinated only (64.7%) or had a prior diagnosis only (0.74%) or both (2.1%) in the past 165 days (hereafter referred to as being immune). At the area level, immunity coverage ranged from 0-63.1% for quintile 1, 63.1%-66.9% for quintile 2, 66.9%-69.2% for quintile 3, 69.2%-72.2% for quintile 4, and 72.3%-100% for quintile 5. Overall, 74.9% of individuals in the highest immunity coverage areas (quintile 5) were immune, compared with 59.6% in the lowest immunity coverage areas (quintile 1) (**Table 1**).

Compared to individuals living in areas with the lowest immunity coverage (quintile 1), those in areas of the highest immunity coverage (quintile 5) had fewer comorbidities and prior hospitalizations, and were less likely to live in rural areas or in neighborhoods characterized by lower income, lower educational attainment, a higher proportion of essential workers, and higher density housing (**Table 1**).

### Immunity and COVID-19 diagnosis

We found evidence of effect modification between individual-level and area-level immunity on COVID-19 diagnosis (p-value<0.001). Higher area-level immunity was consistently associated with lower odds of COVID-19 diagnosis (showing a dose-response relationship) among non-immune individuals (**Figure 2A, Appendix Table 1**). In contrast, among immune individuals, higher area-level immunity was associated with higher odds of COVID-19 diagnosis in the unadjusted model; however, the associations were diminished or reversed after adjustment (**Figure 2A, Appendix Table 1**). After adjustment, the magnitude of the association between area-level immunity (highest vs. lowest) and COVID-19 diagnosis was larger among non-immune individuals (odds ratios [95% confidence interval]: 0.72 [0.70, 0.75]) than among immune individuals (0.93 [0.90, 0.96]) (**Figure 2A, Appendix Table 1**).

**Figure 2.**
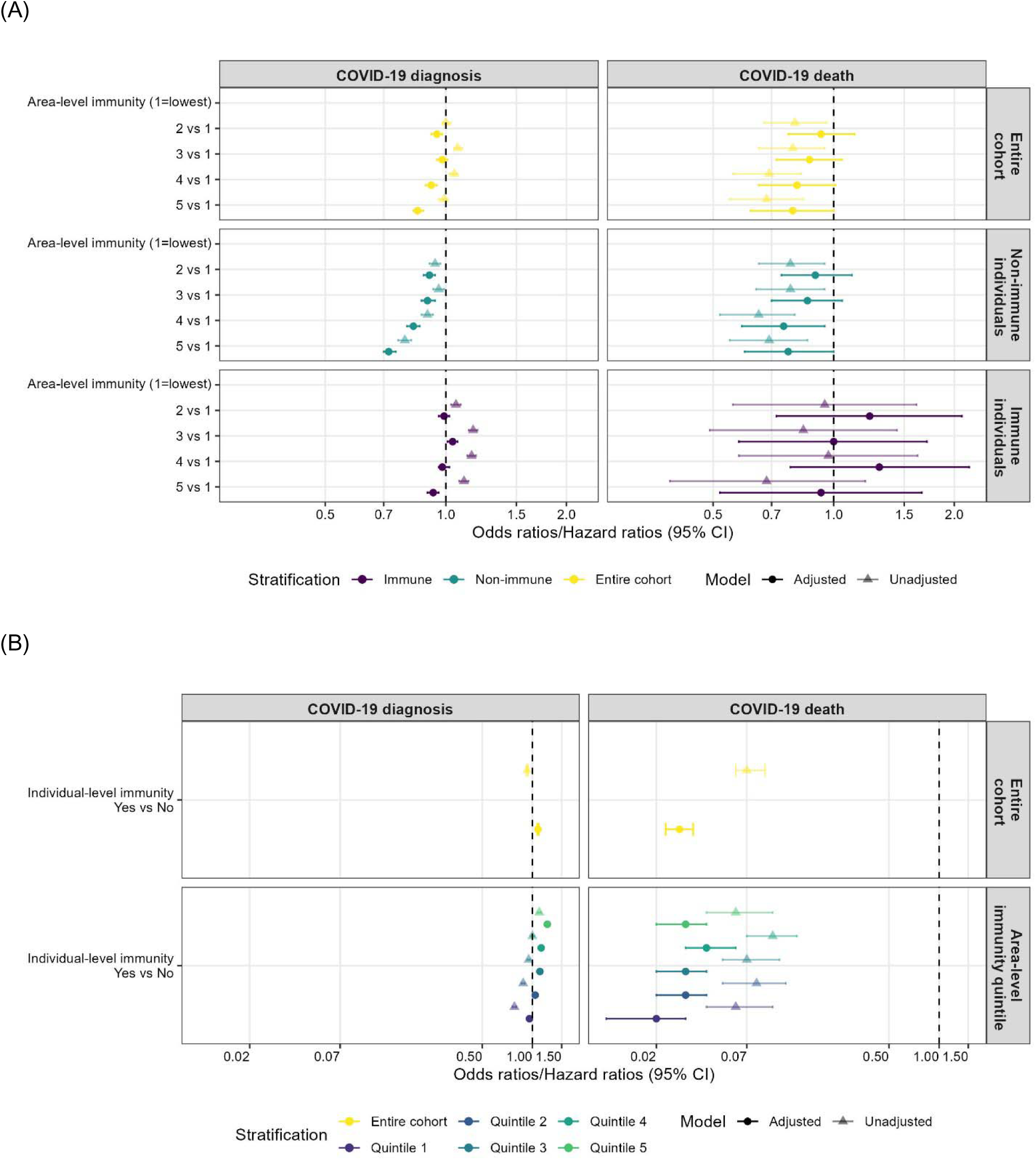
(A) Patterns of area-level immunity in COVID-19 diagnosis and death overall and stratified by individual-level immunity. (B) Patterns of individual-level immunity in COVID-19 diagnosis and death overall and stratified by area-level immunity. Stratified estimates were obtained from models of the entire cohort with interaction terms between individual-level and area-level immunity. Individual-level immunity defined as vaccinated or had prior a positive SARS-CoV-2 test in the past 165 days. Area-level immunity measured by aggregating individual-level immunity at the level of forward sortation area. Adjusted for demographics, baseline health, geography, and area-level social determinants of health.

Similarly, the associations between individual-level immunity and COVID-19 diagnosis varied by area-level immunity. After adjustment, individual-level immunity was associated with lower odds of COVID-19 diagnosis (0.96 [0.93, 0.98]) in the lowest immunity coverage areas but with higher odds of COVID-19 diagnosis (1.23 [1.12, 1.26]) in the highest immunity coverage areas (**Figure 2B, Appendix Table 2**).

### Immunity and COVID-19 death

Higher area-level immunity was also associated with a lower hazard of COVID-19 death among non-immune individuals. For example, the adjusted hazard ratios for COVID-19 death was 0.77 [0.60, 1.00] comparing non-immune individuals in the highest vs. lowest immunity coverage areas (**Figure 2A, Appendix Table 1**). In contrast, we did not find a statistically significant association between area-level immunity and COVID-19 death among immune individuals (**Figure 2A, Appendix Table 1**). However, the statistical test of the interaction term between individual-level and area-level immunity on COVID-19 death was not significant (P-value=0.38), due to a small number of COVID-19 deaths (therefore wide confidence intervals in the estimates) among immune individuals.

Individual-level immunity was consistently associated with a lower hazard of COVID-19 death across different area-level immunity coverage quintiles (**Figure 2B, Appendix Table 2**). Overall, the adjusted hazard ratio for COVID-19 death was 0.03 [0.02, 0.03] comparing immune vs. non-immune individuals.

### Sensitivity analysis

In the sensitivity analyses where area-level immunity was measured at the dissemination area level, the results for COVID-19 diagnosis were very similar to those from the primary analyses. However, for COVID-19 death, the association appeared smaller in magnitude and did not persist after adjustment (**Appendix Figure 1**). Sensitivity analyses using a definition of immunity as only vaccination-acquired yielded similar results as the primary analyses for both outcomes (**Appendix Figure 2**).

## Discussion

Using area-level immunity coverage as a proxy measure of herd immunity, our study provides quantitative observational evidence of indirect herd benefits from vaccination and/or prior infection on SARS-CoV-2 infections and COVID-19 deaths, in a setting with a high overall immunity coverage (67.6%). We found that individuals living in areas with higher area-level immunity experienced a lower risk of COVID-19 diagnosis and death, after accounting for individual-level demographics, baseline health, individual-level immunity, and area-level social determinants of health. We also identified interactions between area-level and individual-level immunity. The protective benefit of area-level immunity was greater among non-immune individuals than immune individuals. Similarly, the protective benefit of individual-level immunity was more pronounced among individuals living in lower immunity coverage areas than those in higher immunity coverage areas.

The mechanism by which herd immunity leads to indirect protective benefits is well established (8) and has been demonstrated with simulation models of transmission dynamics of various infectious diseases, including COVID-19(9,10), influenza(11), and pertussis(26). However, empirical/observational evidence on a vaccine’s indirect herd benefits remains limited. Existing studies have primarily focused on household transmission, demonstrating the indirect benefits of vaccines via reduced infectiousness of vaccinated and of infected individuals, which lowers the risk of transmission to their household contacts(5,6,27–30). The largest of such studies was a retrospective cohort study of more than 155,000 households in Israel, which found that in households with two vaccinated parents, there was a 72% and a 58% decreased risk of SARS-CoV-2 infections among vaccine-ineligible children during the Alpha-dominant and Delta-dominant periods, respectively(27).

A few observational studies have provided some community-level empirical evidence for the indirect herd benefits of vaccines(12,14,31), including one focused on COVID-19 vaccines(31). The study examined COVID-19 vaccination coverage and testing data across 177 regions in Israel and found that for every 20% absolute increase in vaccination coverage, the positive test fraction of the unvaccinated population in the community decreased by approximately half(31). However, given the ecological study design, this study was not able to account for individual-level heterogeneity across communities that could partially explain variations in positive test fractions. Our study builds on the existing evidence by demonstrating the independent effect of herd immunity after accounting for both individual-level and area-level heterogeneity.

Our findings on a less pronounced herd immunity benefit among immune persons than non-immune persons support our hypothesis around immune saturation(8). Once a person gains immunity from vaccination or prior infection, the benefits from herd immunity exhibit diminishing returns. However, it is important to note that there was still an indirect herd benefit of vaccination among immune individuals, particularly in areas with the highest immunity coverage (72.3-100%). Our findings reinforce the importance of achieving high vaccination coverage to protect the unvaccinated, including vulnerable populations who may not be able to get vaccinated due to either access barriers, and/or medical mistrust(32), and to provide additional protection to those already vaccinated, including those who experience disproportionate risks despite vaccination.

Our findings showing greater individual-level vaccine effectiveness in areas with lower immunity coverage suggest that herd immunity may act as an effect modifier in observational studies of vaccine effectiveness. A key assumption in vaccine effectiveness studies is “non-interference”, meaning that vaccination of one individual does not affect the risk of infection of others(33). However, this assumption is violated in the presence of herd immunity(2). Herd immunity modifies an individual’s exposure risk by reducing the number of susceptible individuals in one’s contact network, thereby reducing the overall transmission of the virus within the network. For example, in areas with higher vaccination coverage, the number of potential persons “susceptible” to the virus is reduced, lowering the probability that a person who is infectious will come into contact with a person who is “susceptible”, and vice versa. As a result, the ‘effective’ exposure risks are lowered for both vaccinated and unvaccinated individuals, which could lead to a lower estimated magnitude of vaccine effectiveness in populations with high herd immunity. The influence of herd immunity on vaccine effectiveness estimates is often overlooked, or only discussed as a limitation in observational studies of vaccine effectiveness(33). Our study advances this discussion by explicitly considering and treating herd immunity as a potential effect modifier, allowing us to quantify its influence on vaccine effectiveness estimates. Thus, our findings highlight the importance of considering herd immunity when making inferences about vaccine effectiveness estimates from observational studies.

Our study is subject to some limitations. We used immunity coverage of an individual’s residing forward sortation area as a proxy measure for herd immunity. However, an individual’s social interactions and exposures may extend beyond their residing area (e.g., having contacts outside their local area, e.g., employment-related contacts if their workplace is outside the residing area) and change over time (i.e., dynamic rather than static). Future studies can leverage dynamic network models to compare and inform measures of herd immunity. We could not capture all SARS-CoV-2 infections because not all infected individuals were tested. Testing patterns may differ by vaccination status, such that vaccinated individuals may be more likely to be tested for SARS-CoV-2(34), due to unmeasured confounders such as healthcare engagement (**Figure 1**). Residual confounding related to differential testing by vaccination status might explain the ‘negative vaccine effectiveness’ measures observed in areas with higher herd immunity – a bias that has been extensively discussed in the literature(35,36). We limited our focus to effect modification by area-level immunity when examining individual-level immunity, but there are emerging statistical methods and conceptualizations to capture partial interference, which could comprise future work(33). We also limited our focus to effect modification by individual-level immunity when examining area-level immunity, but there are other factors (both individual-level and network-level) that could modify the area-level herd immunity effects(37). Our study was conducted in a setting during a time when the overall immunity coverage was high (67.6%). Cautions should be exercised when generalizing our findings to low immunity coverage settings. While our findings revealed a dose-response relationship between herd immunity quintiles and COVID-19 outcomes, we suggest this relationship is non-linear. Future studies aimed at estimating the herd immunity threshold could further complement the interpretation of our results.

In conclusion, our study provides quantitative empirical evidence supporting the indirect herd benefits of vaccination or prior infection on SARS-CoV-2 infections and COVID-19 deaths in the community. Our findings reinforce the need for high vaccination coverage to protect both unvaccinated populations and those already vaccinated, while also providing insights for interpreting vaccine effectiveness estimates in observational studies in the context of herd immunity.

## Data Availability

The dataset from this study is held securely in coded form at ICES. While legal data sharing agreements between ICES and data providers (e.g., healthcare organizations and government) prohibit ICES from making the dataset publicly available, access may be granted to those who meet pre-specified criteria for confidential access, available at www.ices.on.ca/DAS. The full dataset creation plan and underlying analytic code are available from the authors upon request, understanding that the computer programs may rely upon coding templates or macros that are unique to ICES and are therefore either inaccessible or may require modification.

## Contribution

LW and SM conceptualized the study. LW developed the conceptual framework and analysis plan. LW and SS conducted the data cleaning. LW conducted the data cleaning and statistical analyses. SS, AS, SDB, BS, HS, JCK, and SM provided critical input into the results interpretation and manuscript preparation. LW wrote the manuscript, with review and edits from all co-authors.

## Acknowledgements and disclaimers

This document also used data adapted from the Statistics Canada Postal Code^OM^ Conversion File, which is based on data licensed from Canada Post Corporation, and/or data adapted from the Ontario Ministry of Health Postal Code Conversion File, which contains data copied under license from ©Canada Post Corporation and Statistics Canada. Adapted from Statistics Canada, Canadian Census 2016. This does not constitute an endorsement by Statistics Canada of this product.

Parts of this material are based on data and/or information compiled and provided by: MOH, Canadian Institute for Health Information (CIHI), Cancer Care Ontario (CCO), Immigration, Refugees and Citizenship Canada (IRCC), Statistics Canada, Ontario Health (OH), and IQVIA Solutions Canada Inc. The analyses, conclusions, opinions and statements expressed herein are solely those of the authors and do not reflect those of the funding or data sources; no endorsement is intended or should be inferred.

We thank IQVIA Solutions Canada Inc. for use of their Drug Information File. The authors are grateful to the 14.7 million Ontario residents without whom this research would be impossible.

